# A target trial emulation study to estimate the causal effect of intravenous iron use during pregnancy and its effect on haematological and birth outcomes in Pakistan

**DOI:** 10.64898/2026.08.29.26361700

**Authors:** N Yazdani, E Oakley, A Khan, Muhammad Farrukh Qazi, S Khakwani, A Sheikh, A Mazhar, Uzma Muhammad Iqbal, J Marquis, Bushra Liaqat, Kaveeta Kumari, Ellen C. Caniglia, Aneeta Hotwani, I Nisar, F Jehan, ER Smith, Z Hoodbhoy

## Abstract

**Background:** Despite several trials on the hematological outcomes of intravenous (IV) iron in pregnancy, only few have examined its effect on birth outcomes. We estimated the causal effect of IV-iron on moderate or severe anaemia and birth outcomes.

**Methods:** Women presenting to routine antenatal care in Pakistan with haemoglobin ≤10 g/dL were eligible for treatment. We used target trial emulation (TTE) methodology to estimate the effect of IV-iron treatment within 14 days of anaemia identification, compared to no treatment, on anaemia status at follow-up. A modified TTE analysis examined birth outcomes at delivery for singleton pregnancies, including birthweight, size-for-gestational-age, and mortality. We conducted a separate TTE for each of five gestational-age periods and pooled the results of each TTE.

**Results:** We screened 3115 pregnancies of which 1715 were eligible for IV-iron; 1043 participants were treated during pregnancy. Those who received IV-iron had half the risk of moderate or severe anaemia in pregnancy compared with no treatment (pooled relative risk (RR) 0.40; 95% confidence interval (CI): 0.27, 0.59). The pooled effect of IV-iron on stillbirth suggested an 83% risk reduction (95% CI 55-94%), and trends were similar for perinatal and neonatal mortality.

**Conclusion:** IV-iron treatment improved haematological status in pregnant women and was associated with a large reduction in stillbirth. Given limited data from randomised trials regarding fetal death and treatment earlier in pregnancy, this study contributes important information to the potential benefit of IV-iron in contexts where anaemia and its sequelae are a major public health problem.

**Key Messages:**

1. Our research question was to assess if intravenous (IV) iron, compared with continued prophylactic oral iron alone, improves haematological and birth outcomes for those diagnosed with moderate or severe anaemia during pregnancy.
2. The use of IV-iron for treatment of anemia in pregnancy during routine antenatal care settings may be more effective than continuing prophylactic dose of oral iron supplementation for improving hematological outcomes and reducing the risk of stillbirth, although there was no difference in the risk of preterm or low birth weight babies.
3. Our study assessed the effect of treatment across the gestational age continuum, measured a comprehensive set of maternal and newborn health outcomes which were not studied in many clinical trials, and used data that reflects real world management of anemia where compliance to oral iron may be poor.

## Background

Anaemia in pregnancy remains a pressing public health concern, affecting an estimated 37% of pregnancies worldwide and contributing substantially to poor maternal and neonatal outcomes [1]. The latest National Nutrition Survey from Pakistan reports that over 40% of pregnant women are anaemic, with nearly half presenting with moderate to severe anaemia [2]. This burden exceeds the World Health Organization’s (WHO) threshold for a public health emergency [3] and reflects structural and nutritional vulnerabilities that increase the risk of adverse outcomes.

National guidelines recommend a daily dose of 60 mg of elemental iron plus 400 µg folic acid to prevent anaemia in all pregnant women, and a higher dose (120 mg of elemental iron plus 400 µg folic acid daily) as first-line treatment once anaemia is diagnosed [4]. Prolonged oral iron therapy is challenging in low-resource settings like Pakistan, where women frequently present late in pregnancy leaving insufficient time for oral therapy to restore haemoglobin before delivery [5,6]. Other barriers include poor adherence linked to gastrointestinal side effects of oral iron supplements [7].

Intravenous (IV) iron offers a rapid, effective alternative for correction of moderate or severe anaemia. Iron sucrose (IS) and ferric carboxymaltose (FCM) are commonly used IV formulations, with emerging evidence favouring FCM for its effect on hemoglobin in substantially fewer visits and improved safety profile [8–10]. The hemtological benefits of IV-iron over oral supplementation are well-established in randomised trials [11]. A 2024 Cochrane review of 11 randomised controlled trials (RCTs) (*n=2935*) found IV-iron produced a greater increase in haemoglobin than oral iron, albeit with substantial heterogeneity across trials [12].

Despite increasingly well-established haematological benefits, evidence of IV-iron on pregnancy outcomes remains limited and inconsistent [12]. The RAPIDIRON trial which provided single-dose FCM to women with moderate to severe anaemia demonstrated a modest reduction in low birthweight (LBW) (RR 0.87; 95%CI 0.75-0.99) [13]. On the contrary, two trials including participants with mild anaemia observed no impact on preterm birth or birthweight [14,15]. IV-iron administration initiated in second trimester had modestly better neonatal outcomes compared with late third trimester treatment, suggesting that both timing and anaemia severity at the time of treatment are key determinants of potential neonatal benefit [13,16,17]. Most trials designed for haematological endpoints are heterogenous in baseline anaemia severity, iron formulation, and intervention timing and are underpowered to examine birth outcomes.

No prior study has quantified the causal effect of IV-iron on haematologic and birth outcomes in the Pakistani context. To address this gap, we conducted a Target Trial Emulation (TTE) study using observational data from a large prospective cohort in peri-urban Karachi. This approach enables robust, ethically sound causal inference while mitigating confounding and immortal time bias thus offering an alternative to clinical trials [18]. We hypothesised that IV-iron would substantially improve haematologic status and reduce adverse birth outcomes, particularly when administered early in pregnancy.

## Methods

### Data

The Pregnancy Risk, Infant Surveillance, and Measurement Alliance (PRISMA) is a prospective, open cohort study implemented in two peri-urban communities in Karachi, Pakistan. Participants are enrolled prior to 20 weeks gestation by ultrasound and followed through 1 year postpartum. Clinical data is collected at five antenatal care (ANC) visits (<20-, 20-, 28-, 32-, and 36-weeks gestation). This analysis includes 3115 pregnant women recruited beginning in September 2022 and followed through six weeks postpartum by April 2025. The cohort study protocol has been previously described [19] and further details about the PRISMA Pakistan cohort have been profiled elsewhere [20].

### Intravenous iron treatment procedures

The PRISMA Pakistan study provides ANC in two facilities that use a community midwife-led model to offer IV-iron treatment for moderate or severe anaemia. Women with haemoglobin ≤10 g/dL were offered treatment. However, not all women received timely treatment, often because they did not stay at or return to the facility for treatment. Prior to May 2023, all IV-iron treatment was IS. Beginning in May 2023, both facilities switched treatment to FCM.

As part of routine ANC, all women are provided the standard regimen of daily oral ferrous sulphate (60 mg elemental iron) combined with folic acid (400 µg), intended to support pregnancy nutrition and prevent anaemia. Thus, the comparison group for our study are those who did not receive any treatment for anaemia but may have continued with their prophylactic dose of daily oral iron.

### Outcomes

We defined two primary haematological outcomes measured at the follow-up visit: continuous haemoglobin (g/dL) and moderate/severe anaemia. The 2024 WHO trimester-specific cutoffs were used to define anaemia; we adjusted haemoglobin levels for smoking status [1, 21].

Additionally, we examined several secondary birth outcomes, including stillbirth (>=20-weeks gestation); early neonatal death (0-6 days of life); perinatal death (stillbirth + early neonatal death); neonatal mortality (0-27 days of life); birthweight (grams); LBW (<2500g); preterm birth (<37 weeks); and moderate preterm birth (<34 weeks). We also examined size-for-gestational-age centile and small-for-gestational-age (SGA) status (<10th percentile), based on the INTERGROWTH standards [22].

### Target trial emulation approach

We applied a TTE methodology to our observational data to attempt to estimate the causal effect of IV-iron treatment [23,24]. We conceptualised a model randomised trial (the “target trial”) for assessing the efficacy of IV-iron treatment [23,24], and then used the observational data to emulate the target trial design. A summary of the target trial design and emulation strategy is presented in **Supplementary Material A**.

The target trial protocol has two eligibility criteria: baseline haemoglobin ≤10 g/dL and no previous IV-iron treatment; eligible participants would be randomly assigned to either IV-iron treatment or the untreated group, with treatment within 14 days. To emulate the trial with observational data, we applied the same inclusion criteria and use inverse probability of treatment weighting (IPTW) based on participant and visit characteristics to emulate randomisation. We followed the same treatment timeline by requiring participants to be treated within 14 days of eligibility to be included in the treatment group. Our primary analysis excluded participants treated “late” (i.e., after 14 days but before the next visit). To account for potential selection bias introduced by excluding this group, we conducted a secondary analysis where individuals treated “late” were included in the comparison group (following the intention-to-treat principle). Follow-up haemoglobin was collected at the subsequent visit within 14-70 days of eligibility for the <20- and 20-week visits and within 14-42 days for the 28- and 32-week visits. For the 36-week visit, follow-up measures were drawn from the 6-week postnatal visit.

We conducted separate gestational age-specific TTEs to compare outcomes for participants treated with IV-iron versus those untreated at each visit. For each TTE, we compared follow-up time visually (histograms) and formally compared mean follow-up time by treatment arm using a two-sample t-test.

Those untreated at a given visit (regardless of eligibility) stayed eligible at the subsequent visit if they presented with moderate/severe anaemia again; those treated were no longer eligible for the next TTE. Therefore, the same individuals may appear in multiple TTEs across the five visits but can only appear once in the treatment arm.

For analysis of birth outcomes, the sample was restricted to singleton pregnancies and the comparison group was restricted to participants who remained untreated throughout pregnancy. We applied a set of visit-specific inverse probability weights (IPW) to balance differences between the included sample (those who are treated on time or never treated) and the excluded sample (those who are eligible but treated late or at a subsequent visit). To ensure that our comparisons were valid, we reviewed each visit sample to identify instances where pregnancy outcomes occurred during the 14-day treatment period. For most visits (<20-, 20-, 28-, and 32-weeks), less than 4% of pregnancies ended during the 14-day treatment window. We did not conduct a TTE for the late-pregnancy visit (36-weeks) because 28% of pregnancies ended during the treatment window.

### Statistical analysis

In addition to IPTW to to emulate randomisation, we also employed inverse probability of censoring weighting (IPCW) to account for differences in outcome missingness. Thirty-eight covariates were considered (see **Supplementary Material B**). Time-varying factors were updated for each visit. We estimated IPTW and IPCW using logistic regression models including maternal age group, household wealth quintile, baseline haemoglobin and any other aforementioned variable identified by backwards stepwise selection process where *P*<0.10. For IPCW models, we also included treatment status.

We used linear regression to derive the mean difference (MD) in haemoglobin, birthweight, and size-for-gestational-age centile and generalised linear models to derive the relative risk of moderate/severe anaemia and binary birth outcomes. All models used visit- and outcome-specific IPTW and IPCW. All estimates are presented with 95% confidence intervals. Models were completed in Stata 18.

For haematological outcomes, we pooled estimates across the four visits where the follow-up haemoglobin was collected during pregnancy using a random effects two-stage meta-analysis (R version 4.5.1). For birth outcomes, we pooled estimates across the four TTE estimates.

## Results

Among the 3115 participants, 445 participants were eligible for the <20-week visit (mean gestational age 13.5 weeks); 740 for the 20-week visit (20.1 weeks); 659 for the 28-week visit (27.8 weeks); 552 for the 32-week visit (32.1 weeks); and 334 for the 36-week visit (35.6 weeks). Figure 1 shows the flow chart of participants included in each visit for haematological outcomes.

**Figure 1.**
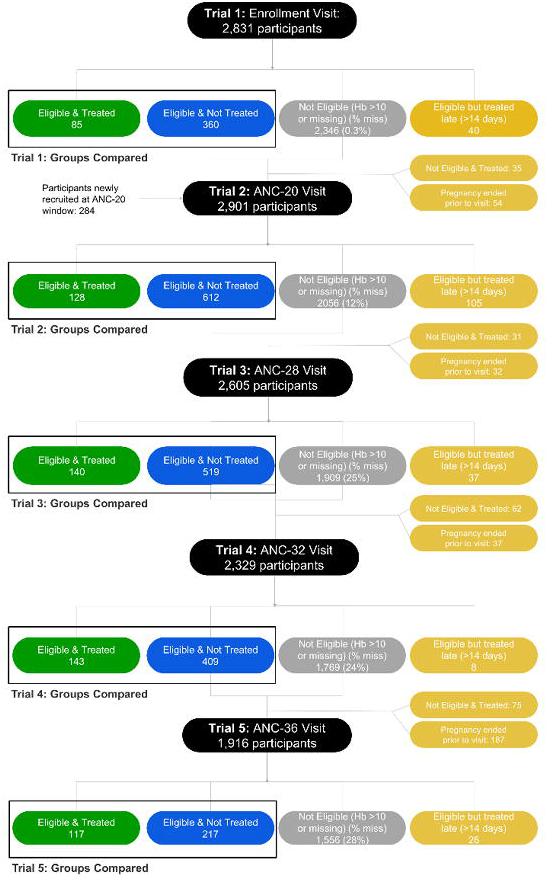
Flow chart of target trial participants for haematological outcomes. Flow diagram tracking participants across five ANC visits; at each visit it shows the numbers eligible and treated with IV-iron within 14 days, eligible but untreated, ineligible or with missing haemoglobin, and those excluded (treated late, treated despite ineligibility, or delivered before the next visit).

Demographic characteristics were similar across treated and untreated groups at all five visits (Table1). Mean participant age was 26-28 years for both groups across visits. Wealth-quintile distribution across both groups were similar, though those treated at the 36-week visit were often in the lowest quantile (30%vs 17%). The treatment group had slightly lower pretreatment haemoglobin.

Those who were untreated were slightly more likely to have a missing haematological outcome at the follow-up visit compared to the treated group at the <20-, 28-, and 36-week visits; this trend was reversed at the 32-week visit and approximately the same at the 20-week visit (see **Supplementary Material C, Table C.1**). Among those with a follow-up measure, we found no difference in mean follow-up time across treatment arms at any visit (see **Supplementary Material C, Table C.2 and Figure C.1**).

In our primary per-protocol analysis, IV-iron raised haemoglobin by 0.97 g/dL compared to the untreated (95% CI: 0.63, 1.31) (Table 2). At the 36-week visit, IV-iron raised haemoglobin by 0.35 g/dL (95% CI: 0.00, 0.70) at 6 weeks postpartum for those treated with IV-iron compared to the untreated.

Across the first four visits, IV-iron more than halved the risk of risk of moderate/severe anaemia at follow-up (pooled RR: 0.40; 95% CI: 0.27, 0.59) versus untreated women (Table 3). At 36 weeks, risk of moderate/severe anaemia at 6 weeks postpartum did not differ between groups.

We repeated this process for the secondary intent-to-treat analysis, finding similar trends in demographics (**Supplementary Material D, Table D1**), outcome missingness (**Table D2**), and mean follow-up time by treatment arm (**Table D3**). In this secondary analysis, we found a similar increase in follow-up haemoglobin (pooled MD 0.89 g/dL [95%CI: 0.61, 1.18], see **Table D4**) and virtually equivalent decrease in moderate-severe anaemia rate (pooled RR 0.38 [95%CI: 0.25, 0.58], see **Table D5**).

### Birth outcomes

After restricting to singletons, 3066 pregnancies were examined in the birth outcome analysis (see flow chart in **Supplementary Material E**). Further details on the demographics and follow-up time are provided in **Supplementary Material F**. Demographics were similar across the treated and untreated groups, with few adverse events occurring within the 14-day treatment window.

Stillbirth risk was lower in the treated group than never-treated group (pooled RR 0.17, 95%CI: 0.06, 0.45), after applying weights related to probability of treatment, censorship, and exclusion due to future treatment. We found no difference for other measures of mortality, including perinatal death (RR 0.39, 95% CI: 0.07, 2.12), early neonatal death (RR 0.90, 95% CI: 0.19, 4.18), or neonatal death (RR 0.52, 95%CI: 0.12, 2.29), or on preterm birth, LBW, and SGA status (Figure 2, **Supplementary Material G, Table G.1**). We found no effect of treatment on pooled birthweight or size-for-gestational-age centile (**Supplementary Material G, Table G.2**).

**Figure 2.**
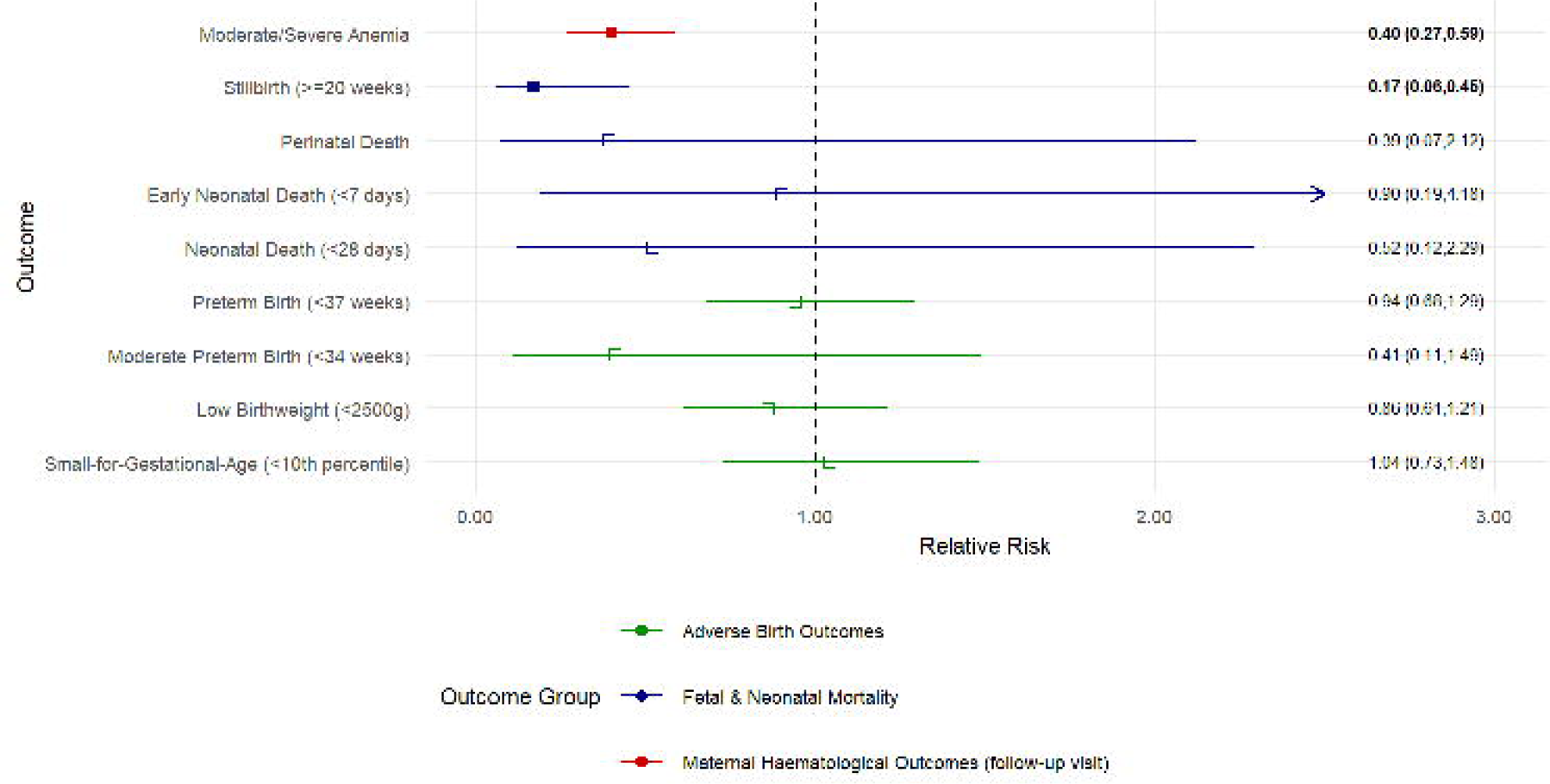
Summary of pooled estimates for maternal haematological and fetal/neonatal outcomes. Forest plot of pooled effect estimates comparing IV-iron with no treatment: mean difference for haemoglobin and relative risks for moderate/severe anaemia, stillbirth, perinatal death, early neonatal death, neonatal death, preterm birth, low birthweight and SGA, each shown with its 95% confidence interval.

## Discussion

Our study demonstrates that midwife-administered IV-iron for anaemic pregnant women, compared with continued prophylactic oral iron alone, during routine antenatal care in Pakistan improved maternal haemoglobin levels and reduced the risk of moderate and severe anaemia when treatment was administered in early pregnancy (<20 weeks gestation) as well as in the third trimester (28 and 32 weeks).We found no difference between IV-iron and oral iron for most adverse birth outcomes though IV-iron may be linked to a stronger reduction in stillbirth risk.

We observed a clinically meaningful increase in haemoglobin (∼1 g/dL) following IV-iron administration, as compared to no treatment. Our study found nearly twice the average effect seen in randomised trials reported in the recent Cochrane Review, though there was a high degree of heterogeneity across studies included in the meta-analysis [12]. Among the 11 trials (*n=2935*) included in the analysis of haemoglobin outcomes [12], the effect of IV-iron compared to oral iron on haemoglobin ranged from a high of 1.04 g/dL (0.54, 1.54 g/dL) [25] to a low of - 0.01 g/dL (-0.44, 0.42 g/dL) [26]. This likely reflects the differences in study design, setting, and inclusion criteria, or residual confounding in our target trial emulation. Most Cochrane trials targeted iron-deficiency anaemia, whereas our treatment decisions rested on haemoglobin alone (iron status was not known at the time of treatment). These trials also spanned wide ranges of gestational age (13–37 weeks) and baseline haemoglobin, which are important modifiers of the intervention effect. Importantly, randomised trials generally compared IV-iron to supervised oral iron, whereas we compared IV-iron to ongoing prophylactic oral-iron; in routine care, oral iron adherence is likely lower, and the prophylactic dose is lower than treatment used in trials. Thus, our findings of a potentially larger benefit is especially important for public health programmes tackling low oral-iron compliance and high anaemia burden.

A recent systematic review and meta-analysis by Pandey and colleagues examined birth outcomes following IV-iron compared to oral iron for the treatment of any anaemia in pregnancy [27]. Consistent with our findings, the systematic review found no effect of IV-iron on birthweight (18 studies, 3735 infants), preterm birth (8 studies, 3738 infants), or neonatal death (3 studies, 2844 infants). However, gestational age at treatment may influence IV-iron’s effect on birth outcomes, and several recent reviews support this hypothesis. The RAPIDIRON trial enrolled participants at 14-17 weeks and found FCM reduced LBW compared with oral iron by 13% (RR 0.87, 95% CI 0.75-0.99) [13]. In contrast, the REVAMP trial enrolled participants throughout the end of the second trimester (13-26 weeks) and found treating anaemia early in pregnancy had no effect on birth outcomes [16]. Similarly, the third trimester REVAMP-TT trial reported no difference in mean birthweights [17], and the IVON trial (20-32 weeks) found no effect on preterm birth (RR 0.94; 95% CI 0.70-1.26) or LBW (RR 0.92 95% 0.60-1.41) [14]. Together, these findings suggest no clear benefit of IV-iron on preterm or low-birth babies, though the effect may depend on treatment timing, baseline anaemia severity, or formulation.

A major contribution of our study was investigating IV-iron’s potential role in reducing stillbirth risk in both early and later pregnancy. Only a few trials have reported on the risk of stillbirth, likely because it is a rare event and trials were generally underpowered to study this outcome. The REVAMP trials in Malawi report no effect of IV-iron treatment in both the trial providing treatment earlier in pregnancy (RR 0.79 95% CI 0.30 to 2.11) [16] and in the trial providing treatment in the third trimester (RR 1.29; 95% CI 0.29 to 5.72) [17]. Similarly, the recent IVON trial in Nigeria (RR 1.12; 95% CI 0.56-2.25) [14] and the Neogi trial in India (0.97; 95% CI 0.52-1.81) [29] reported no effect of IV-iron as compared to oral iron on stillbirth, with wide confidence intervals. In contrast, we found that IV-iron was associated with more than an 80% reduction in stillbirth in current study, with the pooled effect driven by the two target trials including pregnancies treated before 28 weeks gestation. Although our observational results warrant caution, they were consistent across sensitivity analyses aimed at reducing bias. The stronger effect with earlier treatment is biologically plausible: correcting iron deficiency may improve maternal immune function and reduce susceptibility to infections, a recognised contributor to stillbirth risk in late pregnancy.

Our analysis has several strengths. First, TTE methodology with a rich dataset of participant characteristics to derive treatment weights allows us to emulate randomised allocation within the cohort. Sequential haemoglobin measurements throughout pregnancy enabled us to examine treatment effects at multiple gestational age windows. However, several limitations must be considered. The comparator group was prescribed oral iron, but we did not measure adherence. We assume that adherence to oral iron in our population was lower than it would be in a clinical trial, which may have exaggerated the apparent benefit of IV-iron. Despite analytical efforts, residual confounding remains possible. Immortal-time bias cannot be fully excluded; given the lag-time between establishing eligibility and treatment, an outcome may preclude treatment in the untreated group. We imposed a requirement that “treated” participants must have received treatment within 14 days, which limited but may not eliminate potential immortal time bias. Lastly, excluding late treated individuals may limit generalizability, although our intention-to-treat sensitivity analysis, which placed them in the comparator group, found similar effects.

## Conclusion

IV-iron given during routine ANC reduced the risk of moderate and severe anaemia. The observed reduction in stillbirth risk, though requiring cautious interpretation, highlights the potential benefit of identifying and treating anaemia early in pregnancy. Given that FCM can be administered as a single dose, yields greater short-term haemoglobin improvement, and has a more favourable safety profile than older formulations, the entry of lower-cost generic versions represents a new opportunity to expand global access to anemia treatment.

## Data Availability

All data produced in the present study are available upon reasonable request to the authors

## Declarations

### Ethics approval

The study was approved by Aga Khan University Ethics Review Committee: 2022-5920-22763. The study was conducted in accordance with the Declaration of Helsinki.

### Author contributions

Conceptualisation: EO, ERS, ZH, NY, methodology: EO, ERS, ZH, NY, formal analysis: EO, ERS, AM, SK, FQ, NY, ZH; writing – original draft: NY, EO, ZH,ERS writing – review & editing: NY, EO, KA, FQ, SK, AM, UMI, JM, BL, KK, ECC, AH, IN, FJ, ERS, ZH.

ZH is the guarantor for this work.

**Supplementary data** are available at IJE online.

### Data availability

The data underlying this article will be shared on reasonable request to the corresponding author.

### Use of artificial intelligence (AI) tools

The author(s) used ChatGPT Plus to improve the English grammar and readability of this text. The final content was reviewed and approved by the author(s).

