## Supplemental Table _ for "A target trial emulation study to estimate the causal effect of intravenous iron use during pregnancy and its effect on haematological and birth outcomes in Pakistan"

### **Supplementary Material A. Target Trial Protocol and Emulation Strategies**

| **Component** | **Target Trial** | **Emulation** |
| --- | --- | --- |
| Aim | To estimate the effect of IV-Iron treatment for moderate-severe anaemia in pregnant women | Same |
| Eligibility | First-time presentation of moderate to severe anaemia during at ANC visit (<20 weeks, 20 weeks, 28 weeks, 32 weeks, 36 weeks). Women who have been previously treated with IV-Iron are not eligible. | Presentation with moderate to severe anaemia at ANC visit (<20 weeks, 20 weeks, 28 weeks, 32 weeks, 36 weeks); women may or may not have presented with moderate to severe anaemia at a previous visit. Women who have been previously treated with IV-Iron are not eligible. |
| Treatment strategies | Participants are randomly assigned to one of two treatment arms:  1.Receive IV-Iron treatment to treat moderate to severe anaemia within 14 days of moderate/severe anaemia dx  2.No IV-Iron treatment within 14 days of moderate/severe anaemia dx | Patients are non-randomly assigned to IV-Iron at the visit where they are eligible; participants who are treated within 14 days of the eligibility visit are retained in the treatment group. Participants who are not treated at all prior to the relevant outcome (i.e., the follow-up visit for hematological outcomes or delivery for birth outcomes) are considered in the comparison group. Randomization of the treatment is emulated via adjustment for baseline covariates. To account for potential selection bias introduced in the birth outcomes analysis by restricting to those who remain untreated until delivery in the comparison group, we construct an additional set of IPW to these models. |
| Follow-up | Start: Diagnosis of moderate/severe anaemia at ANC visit  End: Follow-up to the next ANC visit or (for the 36-week visit) 6 weeks postpartum | Same |
| Outcome | Anaemia status & continuous haemoglobin at post-randomization follow-up visit (4 weeks post-randomization and 2-4 weeks after treatment of the treated)  Birth outcomes including: stillbirth (at/after 20 weeks gestation); early neonatal death (within the first 7 days of life); perinatal death (stillbirth and early neonatal death); neonatal death (within the first 28 days of life); continuous birthweight; low birthweight status; size-for-gestational-age centile; small for gestational age (<10th percentile); preterm birth (<37 weeks); and moderate preterm birth (<34 weeks). | Anaemia status & continuous haemoglobin at the post-eligibility follow-up visit (up to 10 weeks after initial visit for the <20 week and 20 week visits [70 days]; up to 6 weeks after initial visit for 28 and 32 week visits [42 days]; 6 weeks postpartum for the 36 week visit).  Birth outcomes including: stillbirth (at/after 20 weeks gestation); early neonatal death (within the first 7 days of life); perinatal death (stillbirth and early neonatal death); neonatal death (within the first 28 days of life); continuous birthweight; low birthweight status; size-for-gestational-age centile; small for gestational age (<10th percentile); preterm birth (<37 weeks); and moderate preterm birth (<34 weeks). |
| Causal contrast | Intention-to-treat (ITT) effect, Per-protocol effect | Observational analogs to Per-protocol effect (primary analysis) and ITT effect (secondary analysis) |
| Statistical analysis | Intention-to-treat analysis, Per-protocol analysis | Per-protocol + adjustment for baseline covariates (IPW) (primary analysis)  ITT + adjustment for baseline covariates (IPW) (secondary analysis) |

### **Supplementary Material B. Baseline and visit-specific covariates use to construct inverse probability weights**

| **Covariate** | **Definition** | **Time-varying** |
| --- | --- | --- |
| Demographics | | |
| Age group | Categorical variable of age at enrollment using the following groups: <20 years; 20-34 years; 35+ years. Age categories are required to be included in each IPTW model (regardless of p-value). | No |
| Ethnicity | Categorical variable of ethnicity using the following groups: Punjabi; Bengali; Urdu-speaking; Pashtun; Other (including Seraiki, Balochi, and others) | No |
| Socioeconomic status & household characteristics | | |
| Wealth quintile | We constructed a principal component analysis (PCA) asset index across the entire Pakistan PRISMA pregnancy cohort and assigned each participant to a wealth quintile based on PCA score. Quintiles (1-5) are required to be included in each IPTW model (regardless of p-value). | No |
| Woman’s paid work | Binary indicator of whether the participant reports having any paid work at enrollment. | No |
| Woman’s skilled job | Binary indicator of whether the participant reports having a skilled job at enrollment. | No |
| Woman’s husband has a skilled job | Binary indicator of whether the participant reports having a husband with a skilled job at enrollment. | No |
| Female-headed household | Binary indicator for the head-of-household being a woman (whether the participant herself or another female family member). | No |
| Household rents their dwelling | Binary indicator of whether the household rent their current dwelling at baseline. | No |
| Number of household members per room | Calculated as the number of household members (including children/infants) divided by the number of rooms in the dwelling. | No |
| Traditional stove use | Binary indicator that household cooking is “usually” done on a traditional stove. | No |
| Drinking water source | Binary indicator that the household’s primary drinking water source is piped to their dwelling. | No |
| Improved sanitation | Binary indicator for having improved sanitation at baseline. | No |
| Rudimentary housing materials | Binary indicator for any rudimentary materials used in current dwelling (including for the floor, walls, or roof). Examples of rudimentary materials for housing include dirt floors, cardboard or unfinished plywood in walls or roof, etc. | No |
| Household internet access | Binary indicator for the household having internet access at baseline. | No |
| Smoking inside the home | Binary indicator for the woman herself or any household members smoke tobacco inside the home. | No |
| Woman’s background & agency factors | | |
| Woman’s education | Binary indicator for the woman having any education (1 or more years of schooling) vs. no education (never attended or less than 1 year). | No |
| Woman was married under age 18 | Indicator for the woman experiencing child marriage (i.e., being married under the age of 18). | No |
| Woman has her own mobile phone | Binary indicator for whether the woman has her own mobile phone at baseline. | No |
| Recent use of modern birth control | The woman reported using a modern form of birth control in the 12 months prior to pregnancy. | No |
| Birth location decision | The woman report that she will make the decision about where to give birth alone or together with her partner (compared to those who report that her partner alone or another family member would make the decision) | No |
| Nutritional & health factors | | |
| Early pregnancy BMI category | BMI category (underweight, normal, overweight, obesity) is determined at 9 weeks gestation; for those enrolled after 9 weeks, we use an imputation model to estimate BMI at 9 weeks. | No |
| Health <150cm | Binary indicator for the woman having short stature (<150 cm). | No |
| Substance use | The woman uses any substances, including smoking cigarettes, chewing tobacco, or chewing betelnut. | No |
| Chronic health condition at enrollment | The woman has chronic hypertension or pre-existing diabetes mellitus at enrollment. | No |
| Obstetric history | | |
| Parity | Categorical variable that includes the following groups: para 0; para 1; para 2; para 3; para 4; para 5+ | No |
| History of pregnancy loss | Binary indicator for history of pregnancy loss, including stillbirth or miscarriage. Note that women without previous pregnancy are coded as “0” for this variable. | No |
| Previous pregnancy complication | Binary indicator for any of the following previous pregnancy complications: postpartum hemorrhage; antepartum hemorrhage; preterm birth; prolonged or obstructed labor; cesarean delivery; and/or fetal anomaly. | No |
| Baseline characteristics of current pregnancy | | |
| Facility | Binary indicator for which (of two) centers where the woman sought ANC. | No |
| GA at enrollment | Gestational age at enrollment/first ANC visit (binary indicator for enrolled in first trimester vs. second trimester) | No |
| Wanted current pregnancy | Binary indicator for the woman reported wanting to be pregnant at the time of the current pregnancy | No |
| Assessments & symptoms | | |
| Iron deficiency at enrollment | Binary indicator for iron deficiency at enrollment. Iron deficiency is defined as ferritin <15μg/L or, for those with inflammation, ferritin <70μg/L. | No |
| Depression symptoms | Total score on the Edinburgh Postnatal Depression Scale (EPDS); we rely on the most recent score registered at or before the current ANC visit. | Yes |
| Fatigue symptoms | Fatigue symptoms as reported on selected items from the modified Functional Assessment of Chronic Illness Therapy (FACIT)-Fatigue Scale. We include symptoms in the model as binary indicators where the participant reported each symptom as “quite a bit” or “very much”:   - “I feel fatigued” - “I feel weak all over” - “I feel listless (‘washed out’)” - “I need to sleep during the day” - “I am too tired to eat”   We also include the following symptom, reverse coded:   - “I am able to do my usual activities” | Yes |
| Syphilis infection | Baseline syphilis infection | No |
| Hepatitis infection | Baseline Hepatitis infection (including Hepatitis B or Hepatitis C) | No |
| Visit-specific factors | | |
| Haemoglobin level | Haemoglobin level at the eligibility visit based on complete blood count (CBC), or point-of-care test if CBC is unavailable | Yes |
| Day of the week of visit | Day of the week of the eligibility visit | Yes |
| Form completion ID | Staff member who completed the form for the eligibility visit | Yes |

### **Supplementary Material C.** Primary Analysis (Per Protocol) Supporting Methods Tables

#### Supplementary Table C.1 Comparison of outcome missingness by treatment arm by visit

|  | Treated | Untreated |
| --- | --- | --- |
|  | n/N (%) | n/N (%) |
| <20-week visit | 10 / 85 (11.8%) | 74 / 360 (20.6%) |
| 20-week visit | 24 / 128 (18.8%) | 93 / 612 (15.2%) |
| 28-week visit | 13 / 140 (9.3%) | 82 / 519 (15.8%) |
| 32-week visit | 18 / 143 (12.6%) | 56 / 409 (13.7%) |
| 36-week visit | 3 / 117 (2.6%) | 29 / 217 (13.4%) |

*Notes: This table presents the number and percentage of participants with a missing outcome for each visit by treatment arm.*

#### Supplementary Table C.2 Comparison of follow-up time by treatment arm by visit

|  | Treated | | | Untreated | | | t-test p-value |
| --- | --- | --- | --- | --- | --- | --- | --- |
|  | N | Mean | Stdev | N | Mean | Stdev |  |
| <20-week visit | 75 | 49.1 | 14.0 | 286 | 46.0 | 15.2 | 0.12 |
| 20-week visit | 104 | 55.3 | 8.9 | 519 | 54.3 | 9.3 | 0.31 |
| 28-week visit | 127 | 31.4 | 5.6 | 437 | 31.1 | 6.4 | 0.55 |
| 32-week visit | 127 | 29.2 | 7.8 | 354 | 28.6 | 6.9 | 0.41 |
| 36-week visit | 114 | 73.1 | 15.6 | 188 | 72.3 | 16.9 | 0.66 |

*Notes: This table presents the mean and standard deviation for days between the pre-test haemoglobin measure at the visit of interest and the follow-up post-test haemoglobin measure at the subsequent antenatal care visit (or, the postnatal visit for those eligible at the 36-week antenatal care visit). The p-value presented is drawn from a* *two-sample t-test comparing mean follow-up time between the treated and untreated groups at each visit.*

#### Supplementary Figure C.1 Follow-up time by visit by treatment arm

| Panel 1. <20-week visit follow-up time by treatment arm  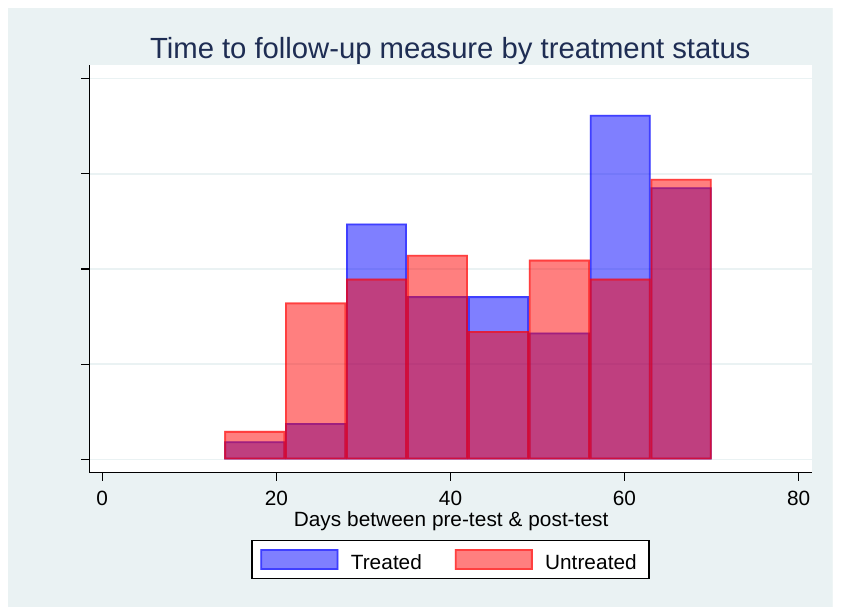 | Panel 2. 20-week visit follow-up time by treatment arm  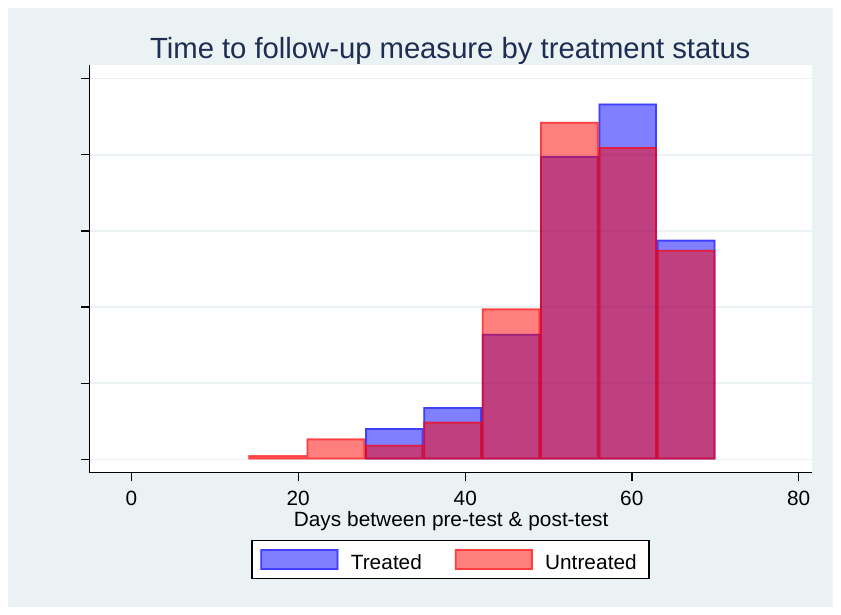 |
| --- | --- |
| Panel 3. 28-week visit follow-up time by treatment arm  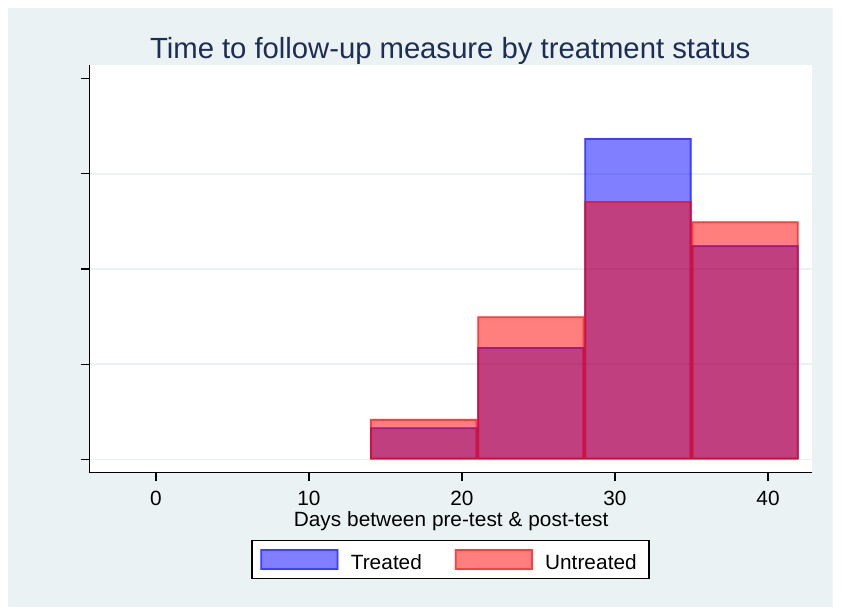 | Panel 4. 32-week visit follow-up time by treatment arm  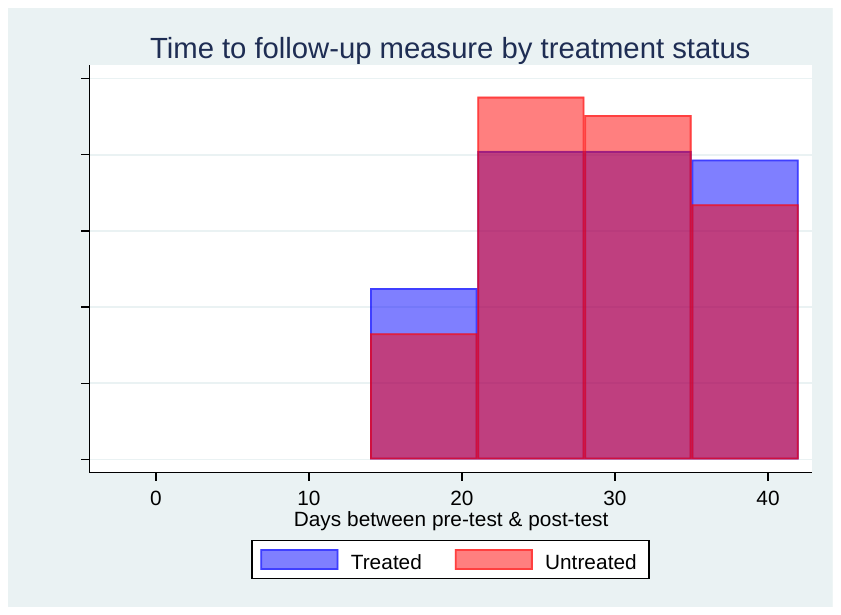 |
| Panel 5. 36-week visit follow-up time by treatment arm  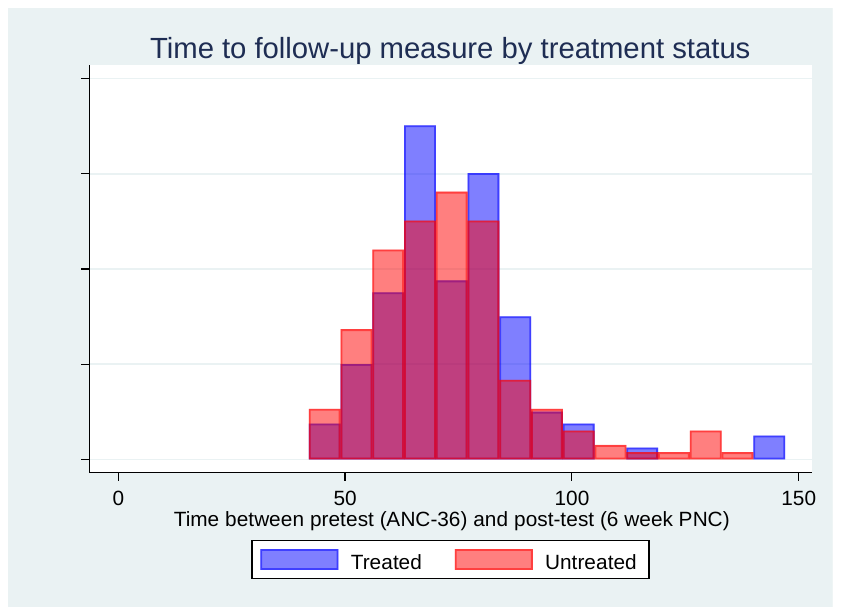 |  |

*Notes: These figures present the distribution of days between the pre-test haemoglobin measure at the visit of interest and the follow-up post-test haemoglobin measure at the subsequent antenatal care visit (or, the postnatal visit for those eligible at the 36-week antenatal care visit).*

### **Supplementary D.** Intent-to-Treat Sensitivity Analysis Demographics, Supporting Methods Tables, and Results

#### **Supplementary Table D1. Demographics by target trial (antenatal care visit) for hematological outcomes**

| **Characteristic** | **Trial 1: <20-week visit** | | **Trial 2: 20-week visit** | | **Trial 3: 28-week visit** | | **Trial 4: 32-week visit** | | **Trial 5: 36-week visit** | |
| --- | --- | --- | --- | --- | --- | --- | --- | --- | --- | --- |
|  | **Treated** | **Untreated** | **Treated** | **Untreated** | **Treated** | **Untreated** | **Treated** | **Untreated** | **Treated** | **Untreated** |
| Total | 85 | 400 | 128 | 717 | 140 | 556 | 143 | 417 | 117 | 243 |
| Mean Age (sd) | 27·4 (6·7) | 27·1 (6·4) | 26·7 (6·0) | 26·9 (6·1) | 27·3 (6·0) | 26·6 (5·9) | 26·6 (5·9) | 26·7 (6·0) | 27·6 (6·0) | 26·3 (5·8) |
| Mean wealth quintile (sd) | 2·6 (1·4) | 2·8 (1·4) | 2·8 (1·5) | 2·8 (1·4) | 2·8 (1·4) | 2·9 (1·5) | 2·8 (1·6) | 3·0 (1·4) | 2·8 (1·4) | 3·1 (1·4) |
| Q1 % | 31% | 27% | 31% | 25% | 26% | 27% | 34% | 22% | 30% | 18% |
| Q2 % | 21% | 16% | 16% | 19% | 17% | 16% | 10% | 17% | 14% | 16% |
| Q3 % | 14% | 18% | 19% | 18% | 22% | 17% | 15% | 20% | 21% | 21% |
| Q4 % | 22% | 23% | 16% | 22% | 24% | 21% | 20% | 22% | 21% | 24% |
| Q5 % | 12% | 16% | 19% | 15% | 11% | 19% | 21% | 18% | 15% | 21% |
| Mean years of education (sd) | 2·8 (4·3) | 1·3 (3·7) | 2·4 (3·6) | 2·5 (3·8) | 2·6 (3·9) | 2·9 (4·0) | 3·1 (4·3) | 2·9 (4·0) | 3·1 (4·3) | 2·6 (3·9) |
| No education % | 68% | 38% | 62% | 66% | 64% | 61% | 59% | 62% | 62% | 63% |
| Para 0 | 38% | 32% | 33% | 29% | 24% | 25% | 21% | 23% | 19% | 24% |
| Para 1-4 | 53% | 55% | 60% | 59% | 61% | 65% | 66% | 65% | 68% | 65% |
| Para 5+ | 9% | 13% | 7% | 12% | 14% | 10% | 13% | 12% | 14% | 11% |
| Mean pretreatment haemoglobin (sd) | 8·3 (1·3) | 9·0 (1·0) | 8·6 (1·0) | 9·2 (0·8) | 8·7 (0·9) | 9·2 (0·7) | 8·9 (0·8) | 9·2 (0·7) | 8·9 (0·8) | 9·3 (0·7) |
| Mean GA (weeks) at pretreatment haemoglobin test (sd) | 13·1 (2·6) | 13·3 (3·1) | 20·1 (1·3) | 20·0 (1·3) | 27·8 (0·9) | 27·7 (1·2) | 31·9 (0·7) | 32·1 (0·8) | 35·5 (1·0) | 35·6 (1·2) |

#### Supplementary Table D.2 Comparison of outcome missingness by treatment arm by visit

|  | Treated | Untreated |
| --- | --- | --- |
|  | n/N (%) | n/N (%) |
| <20-week visit | 10 / 85 (11.8%) | 90 / 400 (22.5%) |
| 20-week visit | 24 / 128 (18.8%) | 109 / 717 (15.2%) |
| 28-week visit | 13 / 140 (9.3%) | 86 / 556 (15.5%) |
| 32-week visit | 18 / 143 (12.6%) | 59 / 417 (14.1%) |
| 36-week visit | 3 / 117 (2.6%) | 33 / 243 (13.6%) |

*Notes: This table presents the number and percentage of participants with a missing outcome for each visit by treatment arm.*

#### Supplementary Table D.3 Comparison of follow-up time by treatment arm by visit

|  | Treated | | | Untreated | | | t-test p-value |
| --- | --- | --- | --- | --- | --- | --- | --- |
|  | N | Mean | Stdev | N | Mean | Stdev |  |
| <20-week visit | 75 | 49.1 | 14.0 | 310 | 46.5 | 14.9 | 0.18 |
| 20-week visit | 104 | 55.3 | 8.9 | 608 | 54.8 | 9.2 | 0.60 |
| 28-week visit | 127 | 31.4 | 0.5 | 470 | 31.3 | 6.4 | 0.86 |
| 32-week visit | 127 | 29.2 | 7.8 | 359 | 28.6 | 7.0 | 0.42 |
| 36-week visit | 114 | 73.1 | 15.6 | 210 | 74.0 | 17.9 | 0.66 |

*Notes: This table presents the mean and standard deviation for days between the pre-test haemoglobin measure at the visit of interest and the follow-up post-test haemoglobin measure at the subsequent antenatal care visit (or, the postnatal visit for those eligible at the 36-week antenatal care visit). The p-value presented is drawn from a two-sample t-test comparing mean follow-up time between the treated and untreated groups at each visit.*

#### Supplementary Figure D.1 Follow-up time by visit by treatment arm

| Panel 1. <20 week visit follow-up time by treatment arm  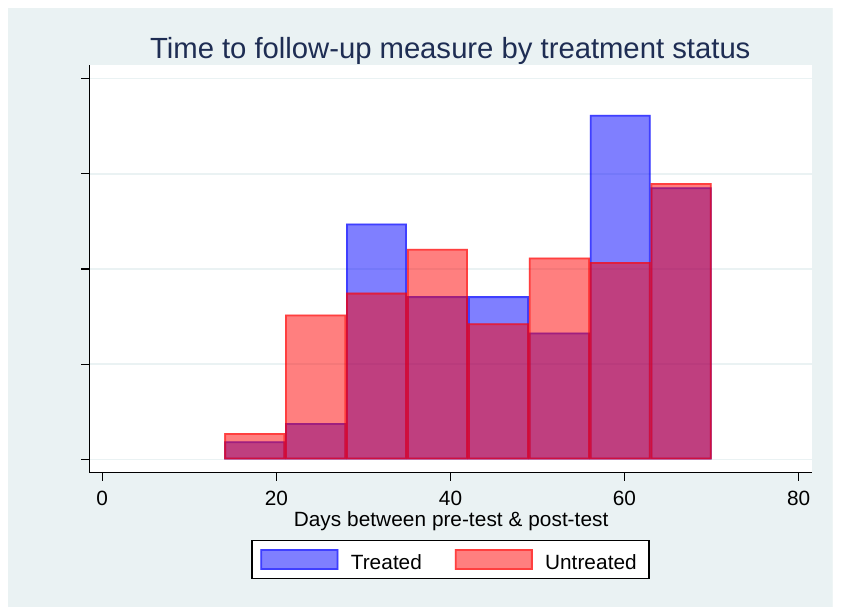 | Panel 2. 20-week visit follow-up time by treatment arm  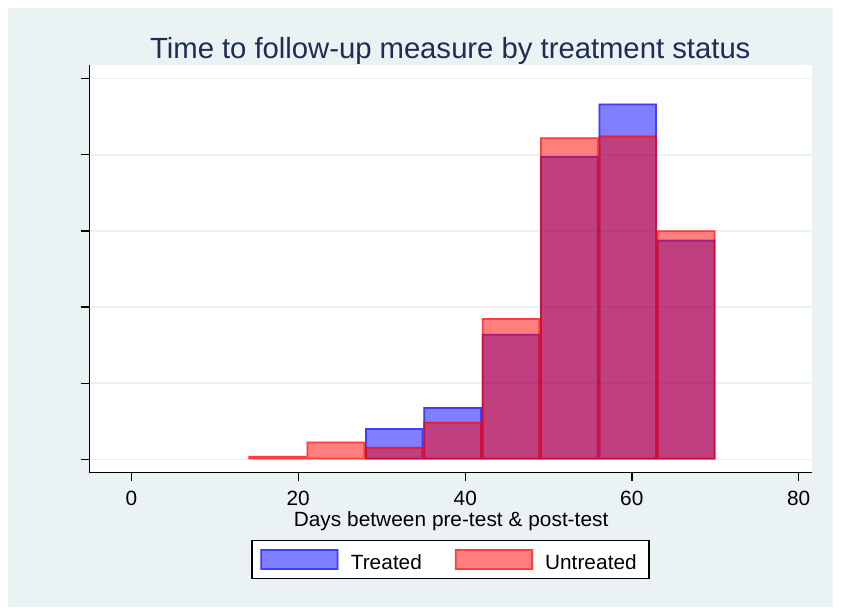 |
| --- | --- |
| Panel 3. 28-week visit follow-up time by treatment arm  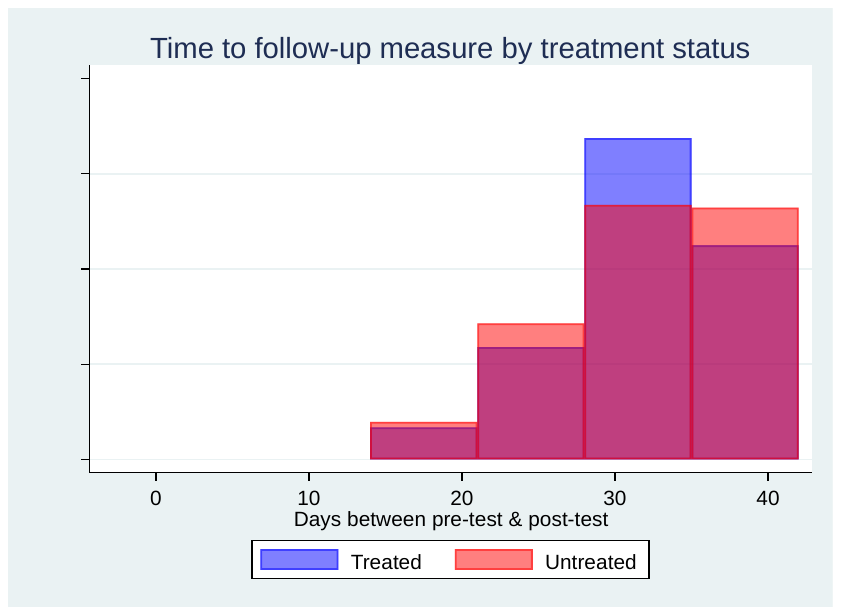 | Panel 4. 32-week visit follow-up time by treatment arm  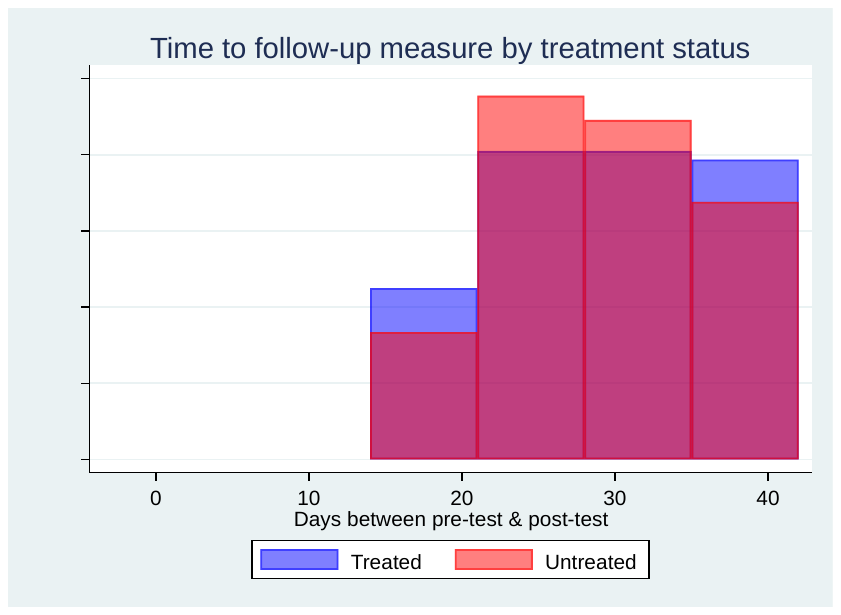 |
| Panel 5. 36-week visit follow-up time by treatment arm  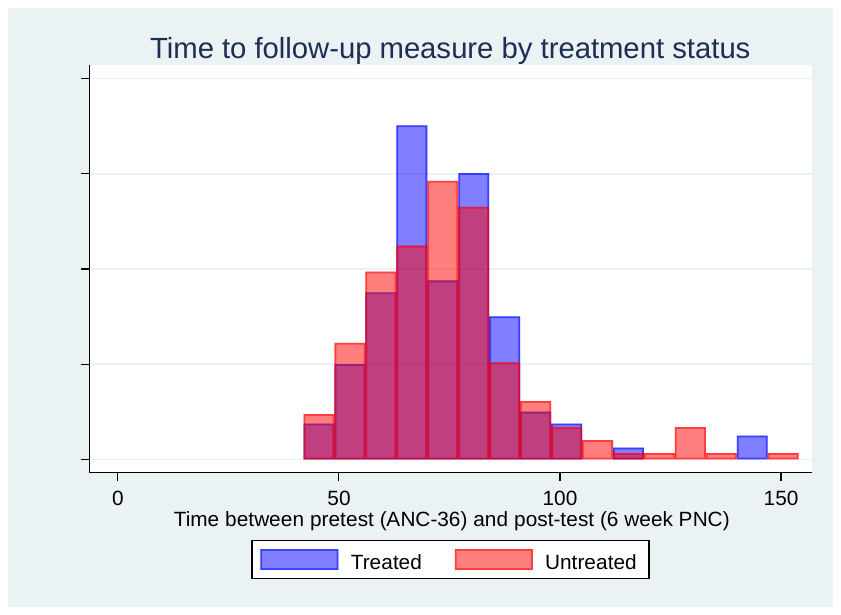 |  |

*Notes: These figures present the distribution of days between the pre-test haemoglobin measure at the visit of interest and the follow-up post-test haemoglobin measure at the subsequent antenatal care visit (or, the postnatal visit for those eligible at the 36-week antenatal care visit).*

#### **Supplementary Table D4· The effect of IV-Iron Treatment on haemoglobin level, stratified by gestational-age specific target trials**

| **Gestational-age specific target trial** | **Sample size (% treated)** | **Mean^1^ haemoglobin in g/dL at Follow-up (SD)** | | **Mean difference^2^ in haemoglobin at follow-up g/dL (95% CI)** |
| --- | --- | --- | --- | --- |
|  |  | **Treated:** | **Untreated:** |  |
| Trial 1: <20-week visit | 485 (18%) | 10·6 (1·4) | 9·3 (1·5) | 1·32 (0·86, 1.78) |
| Trial 2: 20-week visit | 845 (15%) | 10·5 (1·2) | 9·6 (1·3) | 0·63 (0·16, 1·11) |
| Trial 3: 28-week visit | 696 (20%) | 10·2 (1·2) | 9·5 (1·1) | 0·99 (0·70, 1·29) |
| Trial 4: 32-week visit | 560 (26%) | 10·7 (1·4) | 10.2 (1·5) | 0·66 (0·34, 0·98) |
| Pooled Pregnancy Estimate^3^ (<20-weeks, 20-weeks, 28-weeks, 32-weeks) | | | | 0.89 (0.61, 1.18) |
| Trial 5: 36-week visit^4^ | 360 (33%) | 11·9 (1·3) | 11·7 (1·0) | 0·24 (-0·07, 0·56) |

^1^ Treated and untreated means presented are unweighted·

^2^ Mean difference estimates are calculated using linear regression and are weighted based on probability of treatment and probability of censorship (missing outcome) specific to each visit· For each visit, a proportion of follow-up haemoglobin measures are collected after a pregnancy end point (including miscarriage or preterm delivery)· This includes 1% of follow-up observations at <20-weeks, 20-weeks, 28-weeks, and 6% of follow-up observations at 32-weeks.

^3^ Pooled estimates are derived from a DerSimonian-Laird random effects model meta-analysis.

^4^ Results for the 36-week visit are not pooled with the other visit estimates because the follow-up measure for this timepoint is collected postpartum (rather than during scheduled ANC visits).

#### **Table D5· The effect of IV-Iron Treatment on moderate or severe anaemia, stratified by gestational-age specific target trials**

| **Gestational-age specific target trial** | **Sample size (% treated)** | **Events / Total of moderate-severe anaemia at follow-up (%)** | | **Relative risk^1^ of moderate or severe anaemia (95% CI)** |
| --- | --- | --- | --- | --- |
|  |  | **Treated** | **Untreated** |  |
| Trial 1: <20-week visit | 485 (18%) | 13 / 75 (17%) | 169 / 310 (55%) | 0·14 (0·06, 0·31) |
| Trial 2: 20-week visit | 845 (15%) | 24 / 104 (23%) | 327 / 608 (54%) | 0·64 (0·28, 1·45) |
| Trial 3: 28-week visit | 696 (20%) | 51/ 127 (40%) | 321 / 470 (68%) | 0·40 (0·28, 0·58) |
| Trial 4: 32-week visit | 560 (26%) | 34 / 125 (27%) | 176 / 358 (49%) | 0·46 (0·33, 0·64) |
| Pooled Pregnancy Estimate (<20-weeks, 20-weeks, 28-weeks, 32-weeks) | | | | 0·38 (0·25, 0·58) |
| Trial 5: 36-week visit^2^ | 360 (33%) | 19 / 114 (17%) | 45 / 210 (21%) | 0·77 (0·42, 1·41) |

^1^ Relative risk presented for each visit are calculated using generalized linear models and are weighted based on probability of treatment and probability of censorship (missing outcome) specific to each visit. For each visit, a proportion of follow-up haemoglobin measures (and anaemia status derived from the haemoglobin level) are collected after a pregnancy end point (including miscarriage or preterm delivery). This includes 1% of follow-up observations at <20-weeks, 20-weeks, 28-weeks, and 6% of follow-up observations at 32-weeks. Anaemia status for these observations that are collected postpartum are based on the WHO guidelines for non-pregnant adult women of reproductive age (rather than trimester-specific pregnancy cutoffs). Pooled estimates are derived from a DerSimonian-Laird random effects model meta-analysis.

^2^ Results for the 36-week visit are not pooled with the other visit estimates because the follow-up measure for this timepoint is collected postpartum (rather than during scheduled ANC visits).

### **Supplementary Material E. Flow Chart of TTE Participants, restricted to singleton pregnancies (birth outcomes only)**

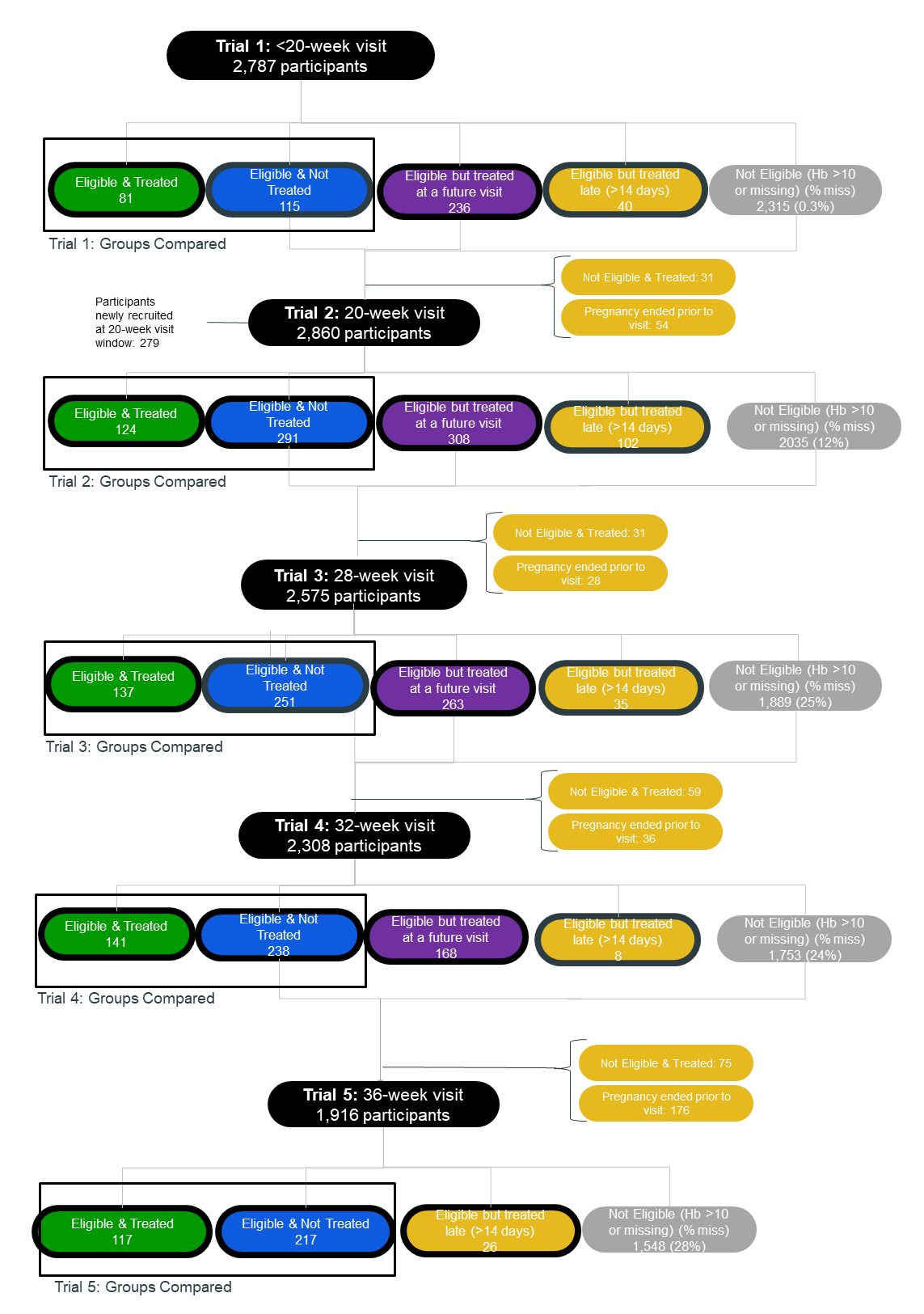

*Note: The birth outcome analysis is restricted to singleton pregnancies and excludes 49 pregnancies with more than 1 fetus (48 sets of twins and 1 set of triplets). Therefore, the total sample size for the birth outcomes analysis is n=3,066 singleton pregnancies. Groups listed in boxes with a* ***bold*** *border are included in the IPW model for likelihood of exclusion due to future treatment.*

### **Supplementary Material F·** Supporting methods tables, birth outcome sample

#### **Table F1·** Demographics by Trial/Visit (birth outcomes only)

| **Characteristic** | **Trial 1: <20-week visit** | | **Trial 2: 20-week visit** | | **Trial 3: 28-week visit** | | **Trial 4: 32-week visit** | |
| --- | --- | --- | --- | --- | --- | --- | --- | --- |
|  | Treated | Untreated | Treated | Untreated | Treated | Untreated | Treated | Untreated |
| Total | 81 | 115 | 124 | 291 | 137 | 251 | 141 | 238 |
| Mean Age (sd) | 27·3 (6·7) | 26·9 (6·8) | 26·6 (6·1) | 26·5 (6·3) | 27·2 (6·0) | 26·4 (5·9) | 26·5 (5·9) | 26·6 (6·3) |
| Mean wealth quintile (sd) | 2·7 (1·5) | 2·9 (1·4) | 2·8 (1·5) | 2·9 (1·4) | 2·8 (1·4) | 2·9 (1·5) | 2·8 (1·6) | 3·1 (1·4) |
| Q1 % | 32% | 23% | 31% | 23% | 25% | 24% | 34% | 21% |
| Q2 % | 19% | 17% | 15% | 21% | 18% | 16% | 11% | 16% |
| Q3 % | 14% | 19% | 19% | 18% | 22% | 15% | 16% | 19% |
| Q4 % | 23% | 23% | 15% | 23% | 25% | 27% | 19% | 25% |
| Q5 % | 12% | 17% | 19% | 15% | 11% | 17% | 21% | 20% |
| Mean years of education (sd) | 2·8 (4·3) | 2·0 (3·6) | 2·5 (3·6) | 2·3 (3·7) | 2·7 (3·9) | 2·7 (3·9) | 3·1 (4·3) | 2·7 (3·9) |
| No education % | 68% | 72% | 61% | 67% | 63% | 62% | 59% | 62% |
| Para 0 | 40% | 37% | 33% | 33% | 25% | 27% | 21% | 23% |
| Para 1-4 | 51% | 49% | 60% | 57% | 61% | 64% | 66% | 65% |
| Para 5+ | 9% | 14% | 7% | 10% | 14% | 9% | 13% | 12% |
| Mean pretreatment haemoglobin (sd) | 8·3 (1·2) | 9·1 (1·1) | 8·6 (1·0) | 9·4 (0·7) | 8·8 (0·9) | 9·4 (0·6) | 8·9 (0·8) | 9·3 (0·7) |
| Mean GA (weeks) at pretreatment haemoglobin test (sd) | 13·1 (2·6) | 13·9 (3·0) | 20·1 (1·3) | 20·1 (1·4) | 27·7 (0·9) | 27·9 (1·2) | 31·9 (0·7) | 32·2 (0·8) |

#### **Table F2·** Summary of pregnancy outcomes and events occurring within the 14-day treatment period by visit

|  | **Trial 1: <20-week visit** | | **Trial 2: 20-week visit** | | **Trial 3: 28-week visit** | | **Trial 4: 32-week visit** | | **Trial 5: 36-week visit^1^** | |
| --- | --- | --- | --- | --- | --- | --- | --- | --- | --- | --- |
|  | Treated | Untreated | Treated | Untreated | Treated | Untreated | Treated | Untreated | Treated | Untreated |
| Total | 81 | 115 | 124 | 291 | 137 | 251 | 141 | 238 | 117 | 217 |
| Mean gestational age (weeks) at pregnancy endpoint (sd) | 37.9 (2.7) | 35.8 (6.6) | 38.0 (2.3) | 37.5 (3.1) | 38.4 (1.7) | 38.0 (2.2) | 38.2 (1.9) | 37.9 (1.8) | 38.7 (1.4) | 38.5 (1.4) |
| Number of pregnancies ending within the 14-day treatment window / Total pregnancies (%) | 0 / 81 (0%) | 3 / 115 (3%)^2^ | 0 / 124 (0%) | 2 / 291 (1%)**^3^** | 0 / 137 (0%) | 1 / 251 (0.4%) | 3 / 141 (2%) | 9 / 238 (4%) | 25 / 117 (21%) | 60 / 217 (28%) |
| Number of adverse events occurring within the 14-day treatment window / Total number of adverse events observed (%) | | | | | | | | | | |
| Stillbirth (>=20 weeks) | 0 / 1 (0%) | 0 / 10 (0%) | 0 / 2 (0%) | 1 / 13 (8%) | 0 / 2 (0%) | 0 / 3 (0%) | 0 / 1 (0%) | 1 / 5 (20%) | 1 / 2 (50%) | 1 / 5 (20%) |
| Neonatal death (<28 days) | 0 / 2 (0%) | 0 / 8 (0%) | 0 / 5 (0%) | 0 / 12 (0%) | 0 / 4 (0%) | 0 / 10 (0%) | 0 / 5 (0%) | 2 / 6 (33%) | 1 / 6 (17%) | 0 / 0 ( - ) |
| Preterm birth (<37 weeks) | 0 / 20 (0%) | 0 / 26 (0%) | 0 / 30 (0%) | 0 / 71 (0%) | 0 / 22 (0%) | 1 / 59 (2%) | 3 / 32 (9%) | 8 / 57 (14%) | 25 / 30 (83%) | 8 / 10 (80%) |
| Low birthweight (<2500g) | 0 / 22 (0%) | 0 / 20 (0%) | 0 / 28 (0%) | 0 / 69 (0%) | 0 / 22 (0%) | 1 / 50 (2%) | 2 / 33 (6%) | 6 / 55 (11%) | 18 / 37 (49%) | 5 / 19 (26%) |
| Small-for-gestational-age (<10^th^ percentile) | 0 / 25 (0%) | 0 / 25 (0%) | 0 / 34 (0%) | 0 / 69 (0%) | 0 / 26 (0%) | 0 / 57 (0%) | 0 / 46 (0%) | 0 / 37 (0%) | 10 / 44 (23%) | 6 / 40 (15%) |

**^1^ Note:** The 36-week visit is excluded from the formal analysis due to a high number of pregnancy outcomes (and adverse events) occurring within the 14-day treatment window.

**^2^ Note:** Although 3 pregnancies end within the 14-day window at the <20-week visit, none of these participants is considered to have an adverse event of interest for this study because all 3 pregnancies end before 20 weeks (i.e., spontaneous abortion). Therefore, these participants are excluded from the denominator for all birth outcomes examined.

**^3^ Note:** Although 2 pregnancies end within the 14-day window at the 20-week visit, one of these two participants is not considered to have an adverse event of interest for this study because the pregnancy ended before 20 weeks (i.e., spontaneous abortion). Therefore, this participant is excluded from the denominator for all birth outcomes examined.

#### Supplementary Table F.3 Comparison of outcome missingness by treatment arm by visit

|  | Treated | Untreated |
| --- | --- | --- |
|  | n/N (%) | n/N (%) |
| **Stillbirth (among all deliveries >=20 weeks)** | | |
| Trial 1: <20-week visit | 0 / 80 (0%) | 0 / 107 (0%) |
| Trial 2: 20-week visit | 0 / 124 (0%) | 0 / 288 (0%) |
| Trial 3: 28-week visit | 0 / 137 (0%) | 0 / 251 (0%) |
| Trial 4: 32-week visit | 0 / 141 (0%) | 0 / 236 (0%) |
| **Neonatal death (among livebirths)** | | |
| Trial 1: <20-week visit | 1 / 79 (1%) | 0 / 97 (0%) |
| Trial 2: 20-week visit | 0 / 122 (0%) | 4 / 275 (1%) |
| Trial 3: 28-week visit | 2 / 135 (1%) | 0 / 248 (0%) |
| Trial 4: 32-week visit | 0 / 140 (0%) | 1 / 231 (0.4%) |
| **Preterm birth (among livebirths)** | | |
| Trial 1: <20-week visit | 0 / 79 (0%) | 0 / 97 (0%) |
| Trial 2: 20-week visit | 0 / 122 (0%) | 0 / 275 (0%) |
| Trial 3: 28-week visit | 0 / 135 (0%) | 0 / 248 (0%) |
| Trial 4: 32-week visit | 0 / 140 (0%) | 0 / 231 (0%) |
| **Low birthweight (among livebirths)** | | |
| Trial 1: <20-week visit | 10 / 79 (13%) | 19 / 97 (20%) |
| Trial 2: 20-week visit | 10 / 122 (8%) | 50 / 275 (18%) |
| Trial 3: 28-week visit | 14 / 135 (10%) | 33 / 248 (13%) |
| Trial 4: 32-week visit | 13 / 140 (9%) | 30 / 231 (13%) |
| **Small-for-gestational-age (among livebirths)** | | |
| Trial 1: <20-week visit | 10 / 79 (13%) | 19 / 97 (20%) |
| Trial 2: 20-week visit | 10 / 122 (8%) | 50 / 275 (18%) |
| Trial 3: 28-week visit | 14 / 135 (10%) | 33 / 248 (13%) |
| Trial 4: 32-week visit | 14 / 140 (10%) | 30 / 231 (13%) |

*Notes: This table presents the number and percentage of participants with a missing outcome for each visit by treatment arm. Note that this analysis does not include IPCW unless missingness reaches at least 3% for one or both treatment arms.*

### **Supplementary Material G·** Pooled Estimates for Birth Outcomes

#### Table G·1· Pooled Estimates for Birth Outcomes (binary)

| **Visit** | **Sample size (% treated)** | **Treated:**  **events / total (%)** | **Untreated:**  **events / total (%)** | **Relative risk of birth outcome (95% CI)** |
| --- | --- | --- | --- | --- |
| Stillbirth (among pregnancies lasting at least 20 weeks) | | | | |
| Trial 1: <20-week visit | 196 (41%) | 1 / 80 (1%) | 10 / 107 (9%) | 0.09 (0.01, 0.73) |
| Trial 2: 20-week visit | 415 (30%) | 2 / 124 (2%) | 13 / 288 (5%) | 0.06 (0.01, 0.44) |
| Trial 3: 28-week visit | 338 (35%) | 2 / 137 (1%) | 3 / 251 (1%) | 0.53 (0.08, 3.46) |
| Trial 4: 32-week visit | 379 (37%) | 1 / 141 (1%) | 5 / 236 (2%) | 0.25 (0.03, 2.19) |
| Pooled Estimate (<20 weeks, 20 weeks, 28 weeks, 32 weeks) | | | | 0.17 (0.06, 0.45) |
| Perinatal Death (among pregnancies lasting at least 20 weeks) | | | | |
| Trial 1: <20-week visit | 196 (41%) | 2 / 80 (3%) | 18 / 107 (17%) | 0.06 (0.01, 0.29) |
| Trial 2: 20-week visit | 415 (30%) | 4 / 124 (3%) | 21 / 284 (7%) | 0.13 (0.02, 0.65) |
| Trial 3: 28-week visit | 338 (35%) | 5 / 136 (4%) | 11 / 251 (4%) | 2.91 (0.64, 13.36) |
| Trial 4: 32-week visit | 379 (37%) | 4 / 141 (3%) | 9 / 235 (4%) | 0.84 (0.22, 3.30) |
| Pooled Estimate (<20 weeks, 20 weeks, 28 weeks, 32 weeks) | | | | 0.39 (0.07, 2.12) |
| Early Neonatal Death (among livebirths) | | | | |
| Trial 1: <20-week visit | 196 (41%) | 1 / 79 (1%) | 8 / 97 (8%) | 0.04 (0.00, 0.36) |
| Trial 2: 20-week visit | 415 (30%) | 2 / 122 (2%) | 8 / 271 (3%) | 0.74 (0.12, 4.20) |
| Trial 3: 28-week visit | 338 (35%) | 3 / 134 (2%) | 8 / 248 (3%) | 4.18 (0.81, 21.69) |
| Trial 4: 32-week visit | 379 (37%) | 3 / 140 (2%) | 4 / 230 (2%) | 1.44 (0.26, 8.00) |
| Pooled Estimate (<20 weeks, 20 weeks, 28 weeks, 32 weeks) | | | | 0.90 (0.19, 4.18) |
| Neonatal Death (among livebirths) | | | | |
| Trial 1: <20-week visit | 196 (41%) | 2 / 78 (3%) | 8 / 97 (8%) | 0.09 (0.01, 0.56) |
| Trial 2: 20-week visit | 415 (30%) | 5 / 122 (4%) | 12 / 271 (4%) | 0.63 (0.15, 2.58) |
| Trial 3: 28-week visit | 338 (35%) | 4 / 133 (3%) | 10 / 248 (4%) | 3.51 (0.75, 16.51) |
| Trial 4: 32-week visit | 379 (37%) | 5 / 140 (4%) | 6 / 230 (3%) | 0.26 (0.04, 1.54) |
| Pooled Estimate (<20 weeks, 20 weeks, 28 weeks, 32 weeks) | | | | 0.52 (0.12,2.29) |
| Preterm Birth (<37 weeks) (among livebirths) | | | | |
| Trial 1: <20-week visit | 196 (41%) | 20 / 79 (25%) | 26 / 97 (27%) | 1.01 (0.49, 2.06) |
| Trial 2: 20-week visit | 415 (30%) | 30 / 122 (25%) | 71 / 275 (26%) | 1.20 (0.68, 2.11) |
| Trial 3: 28-week visit | 338 (35%) | 22 / 135 (16%) | 59 / 248 (24%) | 1.01 (0.48, 2.12) |
| Trial 4: 32-week visit | 379 (37%) | 32 / 140 (23%) | 57 / 231 (25%) | 0.65 (0.36, 1.18) |
| Pooled Estimate (<20 weeks, 20 weeks, 28 weeks, 32 weeks) | | | | 0.94 (0.68, 1.29) |
| Preterm Birth (<34 weeks) (among livebirths) | | | | |
| Trial 1: <20-week visit | 196 (41%) | 1 / 79 (1%) | 7 / 97 (7%) | 0.13 (0.02, 1.17) |
| Trial 2: 20-week visit | 415 (30%) | 6 / 122 (5%) | 13 / 275 (5%) | 0.85 (0.24, 3.10) |
| Trial 3: 28-week visit | 338 (35%) | 1 / 135 (1%) | 12 / 248 (5%) | 1.76 (0.26, 11.97) |
| Trial 4: 32-week visit | 379 (37%) | 3 / 140 (2%) | 4 / 231 (2%) | 0.11 (0.02, 0.76) |
| Pooled Estimate (<20 weeks, 20 weeks, 28 weeks, 32 weeks) | | | | 0.41 (0.11, 1.49) |
| Low Birthweight (<2500g) (among livebirths) | | | | |
| Trial 1: <20-week visit | 196 (41%) | 20 / 69 (29%) | 22 / 78 (28%) | 1.06 (0.47, 2.36) |
| Trial 2: 20-week visit | 415 (30%) | 28 / 112 (25%) | 69 / 225 (31%) | 0.98 (0.52, 1.84) |
| Trial 3: 28-week visit | 338 (35%) | 22 / 121 (18%) | 51 / 215 (24%) | 1.01 (0.44, 2.33) |
| Trial 4: 32-week visit | 379 (37%) | 33 / 127 (26%) | 55 / 201 (27%) | 0.66 (0.38, 1.15) |
| Pooled Estimate (<20 weeks, 20 weeks, 28 weeks, 32 weeks) | | | | 0.86 (0.61, 1.21) |
| SGA (<10th percentile) (among livebirths) | | | | |
| Trial 1: <20-week visit | 196 (41%) | 25 / 69 (36%) | 25 / 78 (32%) | 0.90 (0.43, 1.86) |
| Trial 2: 20-week visit | 415 (30%) | 34 / 112 (30%) | 69 / 225 (31%) | 1.26 (0.69, 2.31) |
| Trial 3: 28-week visit | 338 (35%) | 26 / 121 (21%) | 57 / 215 (27%) | 0.68 (0.39, 1.18) |
| Trial 4: 32-week visit | 379 (37%) | 37 / 126 (29%) | 46 / 201 (23%) | 1.44 (0.84, 2.47) |
| Pooled Estimate (<20 weeks, 20 weeks, 28 weeks, 32 weeks) | | | | 1.04 (0.73, 1.48) |

Notes: Relative risks presented for each visit are calculated using generalized linear models and are weighted based on probability of treatment, probability of censorship (missing outcome), and probability of exclusion due to future treatment, specific to each visit· Non-live-births are excluded from estimates where the outcome denominator is live birth, including: (early) neonatal death; preterm birth; low birthweight; and SGA· Pooled estimates are derived from a DerSimonian-Laird random effects model meta-analysis·

#### Table G·2 Pooled Estimates for Birth Outcomes (continuous)

| **Visit** | **Sample size (% treated)** | **Treated: mean outcome (sd)** | **Untreated: mean outcome (sd)** | **Mean difference in outcome (95% CI)** |
| --- | --- | --- | --- | --- |
| Size-for-gestational-age centile (among livebirths) | | | | |
| Trial 1: <20-week visit | 196 (41%) | 25.8 (26.0) | 29.7 (27.5) | 1.5 (-12.8, 15.7) |
| Trial 2: 20-week visit | 415 (30%) | 26.5 (23.4) | 28.1 (24.9) | -5.3 (-11.5, 0.9) |
| Trial 3: 28-week visit | 338 (35%) | 28.2 (22.4) | 29.7 (24.4) | 3.8 (-6.2, 13.8) |
| Trial 4: 32-week visit | 379 (37%) | 30.0 (26.3) | 29.7 (24.0) | 1.6 (-4.7, 7.9) |
| Pooled Estimate (<20 weeks, 20 weeks, 28 weeks, 32 weeks) | | | | -0.6 (-5.0, 3.7) |
| Birthweight (among livebirths) | | | | |
| Trial 1: <20-week visit | 196 (41%) | 2,692 (412) | 2,751 (505) | -14 (-167, 195) |
| Trial 2: 20-week visit | 415 (30%) | 2,718 (453) | 2,722 (471) | -135 (-322, 53) |
| Trial 3: 28-week visit | 338 (35%) | 2,807 (415) | 2,748 (464) | -26.7 (-296, 243) |
| Trial 4: 32-week visit | 379 (37%) | 2,767 (459) | 2,741 (432) | 190 (-20, 401) |
| Pooled Estimate (<20 weeks, 20 weeks, 28 weeks, 32 weeks) | | | | 0 (-137, 136) |

Notes: Treated and untreated means presented are unweighted; mean difference estimates are calculated using linear regression and are weighted based on probability of treatment, probability of censorship (missing outcome), and probability of exclusion due to future treatment, specific to each visit· Non-live births are excluded from the model for these outcomes. Pooled estimates are derived from a DerSimonian-Laird random effects model meta-analysis·
